# The process of developing and piloting a tool in the Maldives and Zimbabwe for assessing disability inclusion in health systems performance

**DOI:** 10.1101/2024.02.02.24302175

**Authors:** Hannah Kuper, Phyllis Heydt, Shaffa Hameed, Tracey Smythe, Tapiwanashe Kujinga

**Affiliations:** International Centre for Evidence in Disability, London School of Hygiene & Tropical Medicine, UK; Missing Billion Initiative, USA; Division of Physiotherapy, Department of Health and Rehabilitation Sciences, Stellenbosch University, Cape Town, South Africa; Pan-African Treatment Access Movement, Zimbabwe

**Author notes:** Corresponding author: Hannah Kuper, International Centre for Evidence in Disability, London School of Hygiene & Tropical Medicine, Keppel Street, London WC1E 7HT, UK.

**Keywords:** Disability, health system, assessment

## Abstract

There are 1.3 billion people with disabilities globally. On average, they experience greater healthcare needs and more barriers accessing healthcare. Yet, health systems have failed to adequately include people with disabilities. The purpose of this study was to develop and pilot-test a tool for assessing disability inclusion in health system performance. We presented the “Missing Billion” disability-inclusive health system framework, which includes 4 system-level components and 5 service delivery components, and outputs and outcomes. We developed a tool, consisting of 48 indicators related to the framework components. We consulted international experts, who considered the framework and indicator set to be logical and comprehensive. The tool was pilot-tested in the Maldives (2020) and Zimbabwe (2021), working with local researchers to collect relevant data through document review and key informant interviews. The pilot data demonstrated that collecting data on the indicators was feasible. The tool highlighted areas where the health systems were performing well in terms of disability inclusion (e.g. governance) and other areas where there were large gaps (e.g. leadership) or lack of data (e.g. accessibility, outputs and outcomes). The indicators were updated and refined. We established a process for undertaking the assessment, highlighting the importance of leadership and ownership by the Ministry of Health, to facilitate data collection and implementation of recommendations. In conclusion, this new tool for assessing disability inclusion in health systems performance can help to identify key issues and guide and monitor action.

**Highlights:**

– There are 1.3 billion people with disabilities globally, and they often have poorer health and worse healthcare access than others in the population.
– There is currently no comprehensive tool to assess how inclusive the healthcare system is for people with disabilities. The “Missing Billion” disability inclusive health system conceptual framework was proposed, together with 48 corresponding indicators. The indicator set allows description of the level of disability-inclusion in health systems.
– The indicators were pilot-tested in the Maldives and Zimbabwe and were able to highlight areas of good practice, and identify where further improvements are needed. Some modifications were needed to the indicator set.
– This new assessment approach can help policy makers, in particular at Ministries of Health, to identify key issues and guide action, and thereby may ultimately improve health systems for all.

## 1. Introduction

Globally, there are approximately 1.3 billion people with disabilities (WHO, 2022). People with disabilities experience 15-20 year shorter life expectancy and worse health, on average, than others in the population, because of their underlying impairment and health condition, as well as their frequently poorer and more disadvantaged position in society (Kuper & Heydt, 2019; World Health Organization. World Health, 2011). People with disabilities also often have difficulties receiving appropriate, affordable and quality healthcare due to wide-ranging barriers (e.g. attitudinal, informational and logistical) (Eide et al., 2015; Hashemi et al., 2020; Kuper & Heydt, 2019; WHO, 2022; WHO, 2011). Poorer health and healthcare access is an important issue for the individuals affected and their families, as it impedes their ability to thrive and survive (WHO, 2011). It is also a violation of their fundamental rights - such as the right to the “highest attainable standard of health without discrimination on the basis of disability” as set out in the UN Convention of the Rights of Persons with Disabilities {UN, 2006 #7}. Furthermore, failure to ensure the inclusion of people with disabilities in healthcare will mean that global targets will be difficult to achieve, such as <u>Universal</u> Health Coverage (UHC) and Sustainable Development Goal 3 to “ensure healthy lives and promote well-being <u>for all</u> at all ages” (emphasis added) ({UN, 2015 #600}, Kuper & Hanefeld, 2018; Kuper et al., 2022).

International political commitments have consequently been made to providing disability-inclusive health (UN General Assembly, 2019; WHO, 2021a). For instance, the 2023 Political Declaration of the High-Level meeting on UHC commits to “Ensure availability of and access to health services for all persons with disabilities” {Assembly, 2023 #2257}. Concrete action is now needed to strengthen health systems to improve inclusion of people with disabilities.

For the purpose of this paper, the health system “consists of all organizations, people and actions whose primary intent is to promote, restore or maintain health” (WHO, 2007). Health system performance assessments allow description of the health system to help guide reform and improvement {Papanicolas, 2022 #2262}. A broad variety of tools exist {Papanicolas, 2022 #2262}. However, to date, there is no tool to holistically assess the performance of health systems for people with disabilities. Tools are available to assess the performance of specific components of the health system with respect to disability, such as physical accessibility of facilities (Groenewegen et al., 2021), disability-inclusion in the legal framework (Waddington, 2021), inclusion of people with disabilities in HIV strategies (Hanass-Hancock et al., 2011), or availability of rehabilitation services (Kleinitz et al., 2022). Tools are also available that focus on specific impairment types, such as eye health {Bozzani, 2014 #2258}. There is also a vast literature showing the poorer healthcare experience of people with disabilities (Kuper & Heydt, 2019; WHO, 2022; WHO, 2011), and the lower responsiveness of health systems to their needs {Kibet, 2023 #2259}{Almeida, 2017 #2260}. There is therefore a need to develop a health system performance assessment to measure health systems in terms of disability-inclusion. The new disability-inclusion indicator set would serve three purposes. First, it could be used to help countries, in particular Ministries of Health (MoH), understand how well their healthcare system are performing with respect to the inclusion of people with disabilities and identify areas where more action is needed. Second, repeated use would allow monitoring trends over time and potentially the assessment of the impact of specific interventions (e.g. a new health workforce training on disability, or new policy). Third, international consistent use of the indicators would highlight areas of good practice that could be implemented in other settings.

The aim of this paper is to describe the development of a health system performance assessment tool set to measure the level of disability-inclusion of a health system, and to pilot-test it in the Maldives and Zimbabwe.

## 2. Material and methods

### 2.1 The Missing Billion Framework for a disability-inclusive health system

The Missing Billion framework of a disability-inclusive health system was put forward by two authors (HK and PH), based upon the WHO’s building blocks framework for “strengthening health systems to improve health outcomes”(WHO, 2007), and the Primary Health Care Performance Initiative (PHCPI) Conceptual Framework (Veillard et al., 2017). These frameworks were chosen because they are widely used and were considered potentially appropriate to assess the performance of the health system in terms of disability-inclusion. The WHO’s Building Blocks framework describes six building blocks needed for a strong health system: service delivery; health workforce; information; medical products, vaccines and technologies; financing; leadership and governance (WHO, 2007). The PHCPI framework uses many of the same components as the Building Blocks framework, but emphasises the relationship between, and hierarchy of, the different components (e,g, health financing influences the health workforce and access to drugs and supplies) (PHCPI, 2018; Veillard et al., 2017). It also introduces a greater focus on outputs and outcomes and the “Social determinants and context”. The frameworks were adapted to focus on people with disabilities, by including a further focus on the patient perspective given the specific and additional difficulties that people with disabilities face in accessing healthcare (WHO, 2022), including additional components in relation to disability-specific aspects of the health system (e.g. access to assistive technology - AT, existence of laws protection the right to healthcare of people with disabilities), and reducing some of the details on other components (e.g. PHCPI supportive management: formal training).

The Missing Billion Disability-Inclusive Health System Framework includes 4 system-level components and 5 service delivery components (2 on the demand side, 3 on the supply side) (Figure 1, Table 1). The goal of the framework is to reduce the life expectancy gap experienced by people with disabilities by improving the performance of health systems for people with disabilities. Improving performance with respect to disability-inclusion in these 9 components is hypothesised to improve the output of this system (i.e. “Effective service coverage”) and thereby lead to the ultimate outcome of improved “health status” for people with disabilities.

**Figure 1:**
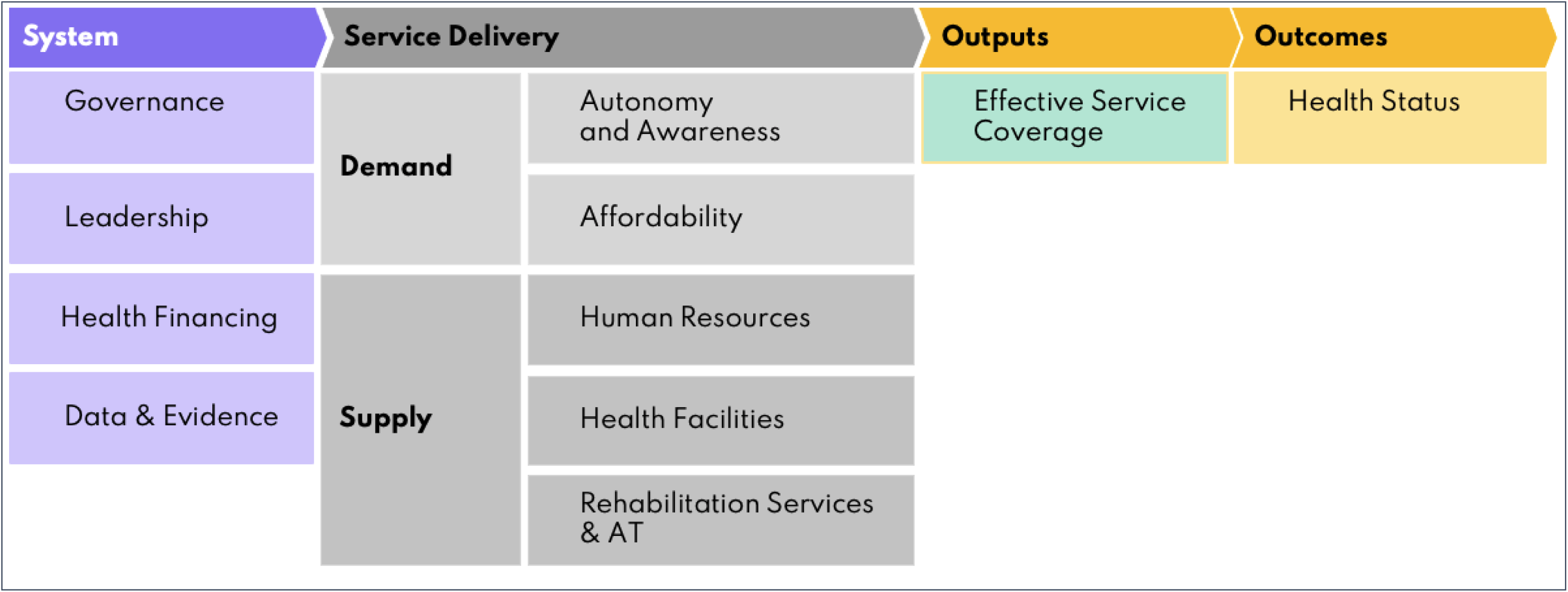
The Missing Billion Health System Framework.

**Table 1:** Definition of the Missing Billion Disability-Inclusive Framework health system components.

| Health system component | Objective |
| --- | --- |
| System |  |
| - Governance | The ratification of international regulations is matched with the creation and adoption of appropriate in-country laws and policies that protect the right to health care for people with disabilities and outlaw discrimination based on disability. Governance mechanisms, particularly those that enable transparency and accountability, must be put in place to enforce this right. |
| - Leadership | Issues around disability are clearly articulated and represented in the MoH, health sector structures and coordination mechanisms. Leadership capacities and structures should be built, strengthened and effectively activated during times of crisis or disasters. |
| - Health financing | Resources are available to ensure health services are accessible to, and affordable for, people with disabilities (e.g. accessibility of facilities and costed reasonable accommodations) and disability-relevant services are provided (e.g. AT). |
| - Data | Routine data showing the health situation of people with disabilities and how their health can be improved are available, and evidence is generated to understand and improve delivery of health services. |
| Service: demand |  |
| - Autonomy and awareness | People with disabilities make their own decisions about health care and are aware of their rights and options. |
| - Affordability | People with disabilities are able to afford health services. |
| Service: supply |  |
| - Human resources | Health-care workforce is knowledgeable about disability and has the skills and flexibility to provide quality care to people with disabilities. |
| - Health facilities | Health-care services, including health-care facility infrastructure and information, are accessible for people with disabilities |
| - Specialized services and AT | Specialized services (e.g. physiotherapy, ophthalmology) and AT are available, affordable and of good quality for people with disabilities. |

The framework was presented to a range of experts through a series of individual meetings, including governmental stakeholders (Georgia, Ireland, Malawi, Ukraine, Poland), UN officials, health systems experts (PHCPI and Ariadne Lab), academics specialising in disability studies, and disability rights organizations (European Disability Forum). The sample included people with disabilities. These key informants were asked about the relevance of the nine components for creating disability-inclusive health systems, and suggestions were sought for improvements.

### 2.2 Development of indicators for a health system performance assessment tool on disability inclusion health system

Two authors (an epidemiologist – HK – and a global health system expert - PH) developed specific indicators to measure the level of disability-inclusion for each framework component.

First, we reviewed indicators proposed to monitor the implementation of article 25 (health) of the UN Convention on the Rights of Persons with Disabilities (UN, 2020), PHCPI and UHC (Veillard et al., 2017; WHO, 2017). We also reviewed a range of toolkits relevant to health system strengthening for people with disabilities to identify possible application and note good practice examples: Strategizing health in the 21^st^ century: a handbook (WHO, 2016), Toolkit frontline health (unpublished), Essential Management Package for Strengthening Physical Rehabilitation Centers (MSH, 2015), Autism toolkit for GPs (no longer available online), Learning Disability Annual Health Check Toolkit (NHS, 2021) and the rapid Assistive Technology Assessment (rATA) (WHO, 2019).

Next, we formulated 3-5 indicators per framework component; we aimed to increase the feasibility of data collection through limiting the number of indicators. We focussed on developing indicators where data would be available, based on our previous experience and internet searches (e.g. of policy documents).

The preliminary indicator set was reviewed by a lead academic specialising in collecting data on disability and a UN representative working on disability. They gave specific feedback on the perceived relevance of the indicator, wording of the indicators and feasibility of collecting the data. We also asked additional key informants about specific areas of assessment and/or sources of information within the context of the indicators. The indicators were adapted, based on the feedback obtained. In total, 34 indicators were developed to measure performance across the 9 health system components, as well as 14 for the output/outcome components.

### 2.3 Pilot-test in Maldives and Zimbabwe

Research partners were selected to pilot test the application of the indicator set in the Maldives (2020) and Zimbabwe (2021) to assess its feasibility. These settings were chosen as they are contrasting in nature and the context of ongoing research of our group, so that there were research partnerships and activities in place. The Maldivian research partner (SH) is a policy specialist, who led a national disability study in the country, which included a national survey, qualitative fieldwork and policy analysis. She also works closely with stakeholders from the MoH and disability groups. The Zimbabwean research partners (TK and TS) are academics who work closely within the Zimbabwean health, policy and legal system. In both settings, the health system performance assessment was undertaken within the context of a broader research study on disability.

The research partners aimed to collect data for all the indicators through reviewing policy documents and internet search of grey literature (e.g. NGO or OPD reports, national policy/strategy reports, laws, ministry of health reports), searching scientific publications and interviewing 3-5 key stakeholders per country (e.g. MoH representatives). They noted whether the data was available, and if so what it showed for each of the indicators, on a standardised excel sheet. They also reported which indicators could not be completed and why, and which indicators need to be re-worded or otherwise clarified, or eliminated. Overall, the researchers provided their qualitative impression on the feasibility of undertaking the assessment. The results were discussed as a group, including consideration of how the indicator set needed to be adapted.

### 2.4 Indicator set and assessment approach refinement

Individual interviews were held with seven health system stakeholders (MoH representatives, NGOs, academics) from a range of countries (South Africa, Bangladesh, Nigeria, Malawi, Indonesia, Uganda, India). The key informants were asked about the perceived value of the indicator set, potential users, challenges to implementation and strategies for encouraging the uptake of findings. A workshop was then held with three international stakeholders and three human-centred design experts to agree recommendations on the use of the indicator set and its scale up.

### 2.5 Ethics

Ethical approval for the study was granted from the Medical Research Council of Zimbabwe (MRCZ), the Maldives National Bureau of Statistics, the National Health Research Committee at the Maldives MoH and the Institutional Review Board of London School of Hygiene & Tropical Medicine, UK.

## 3. Results

### 3.1 Framework indicators

Overall, the research partners in the Maldives and Zimbabwe reported that it was feasible to collect most of the indicators, and that the majority were clearly defined and important. The specific changes suggested for the indicators are described.

#### 3.2.1. Systems level indicators

##### 3.2.1.1. Governance

Four indicators relate to governance (Table 2). The indicator on the ratification of the UN Convention on the Rights of Persons with Disabilities (UNCRPD) was considered to be clear (1.1) (UN, 2006). It was recommended that indicator 1.2 was separated into two distinct indicators as National laws on health differ in their influence to national policies. Description of “inclusion” of people with disabilities across indicators 1.2-1.4 was perceived to be vague and did not specify the quality of the inclusion. The information for these indicators could be collected through an Internet search.

**Table 2:**
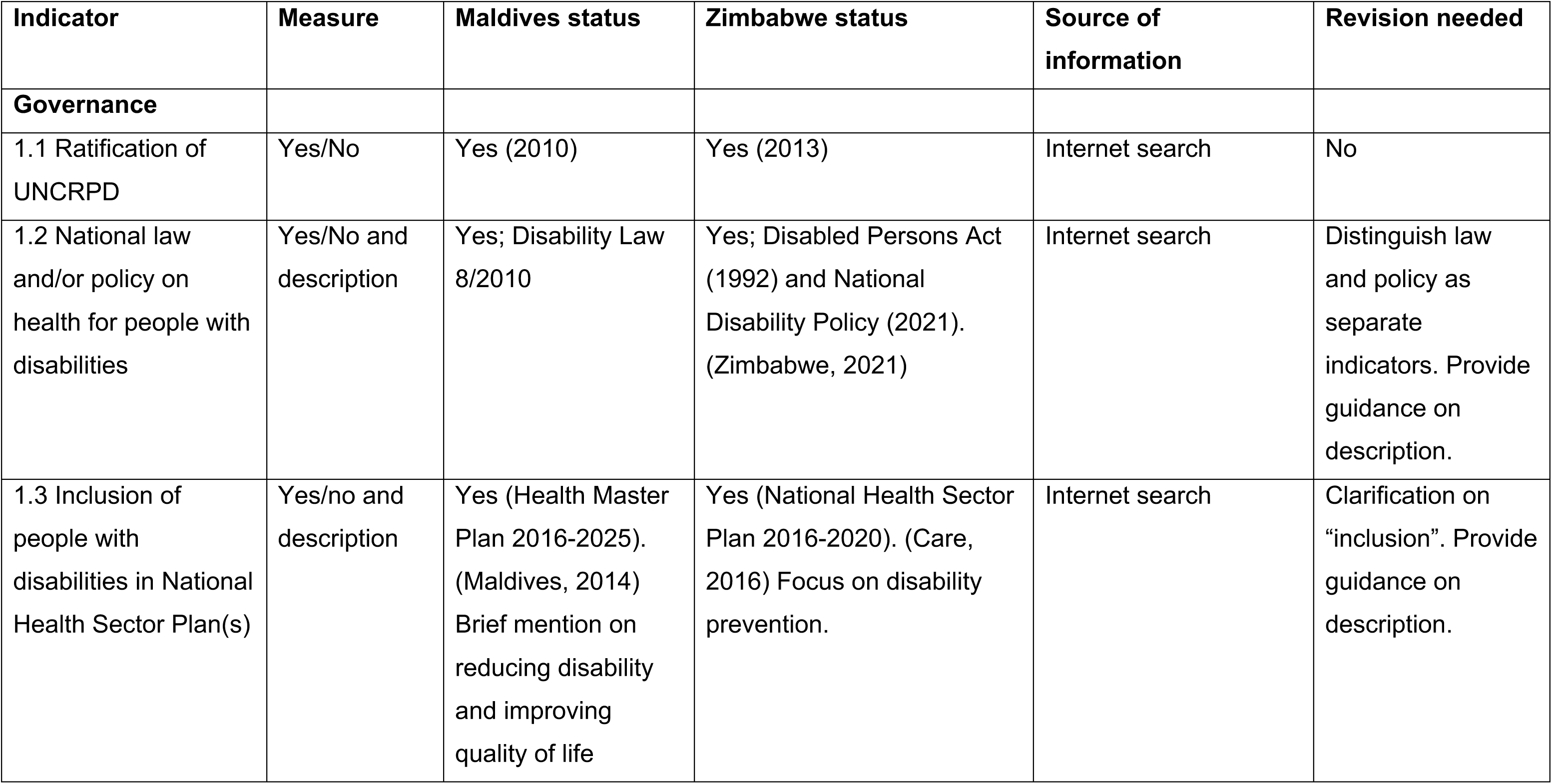
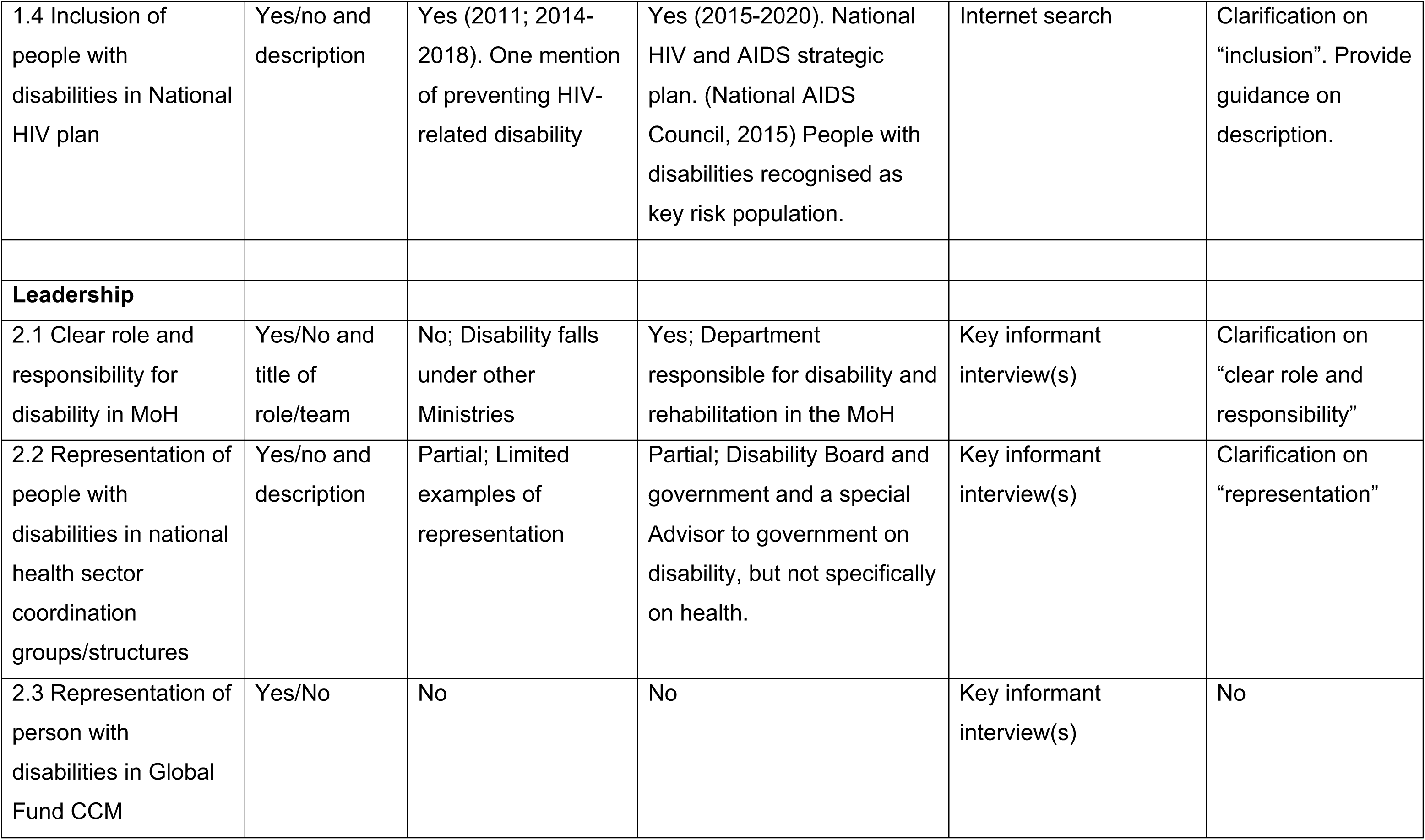
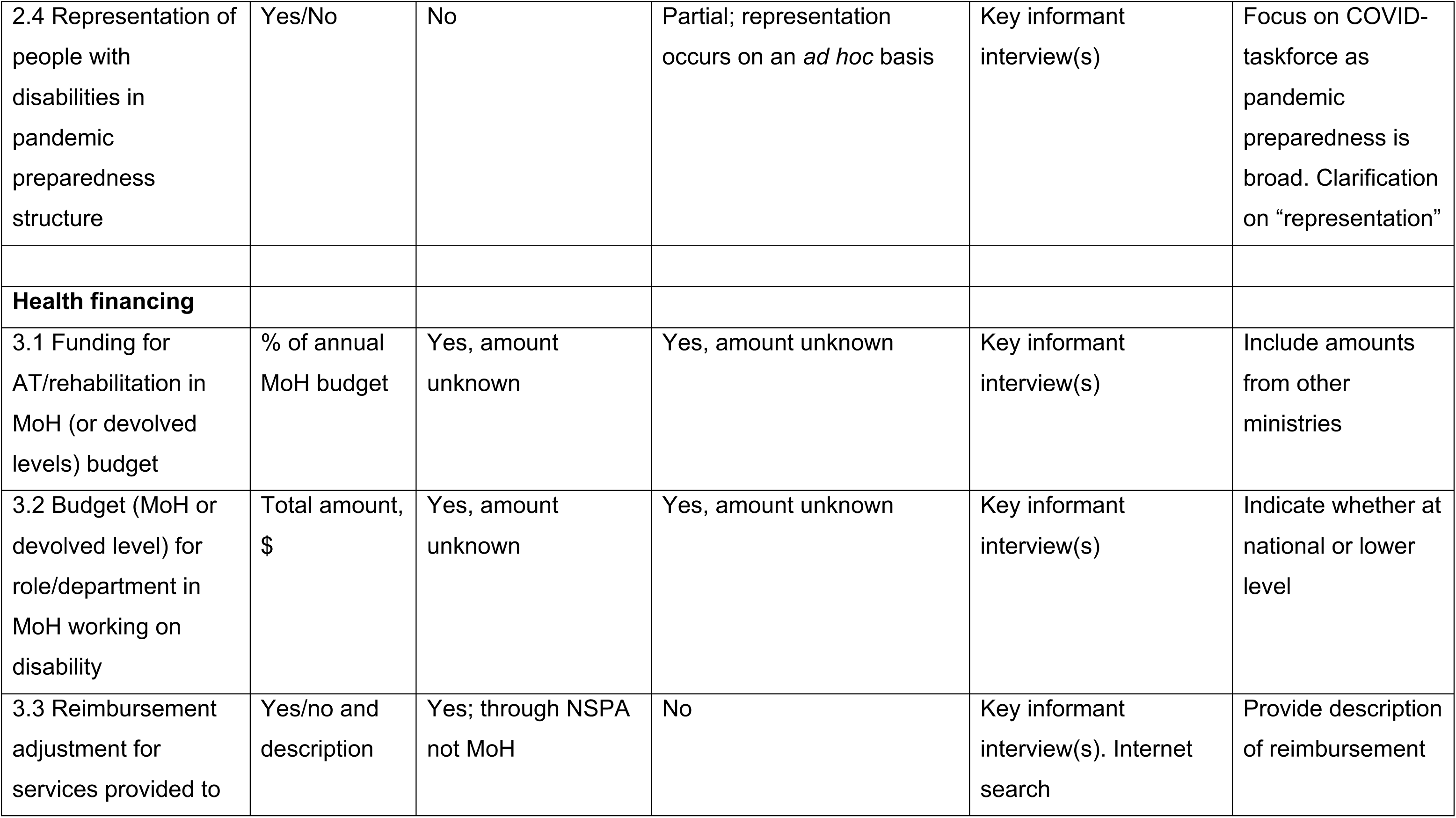
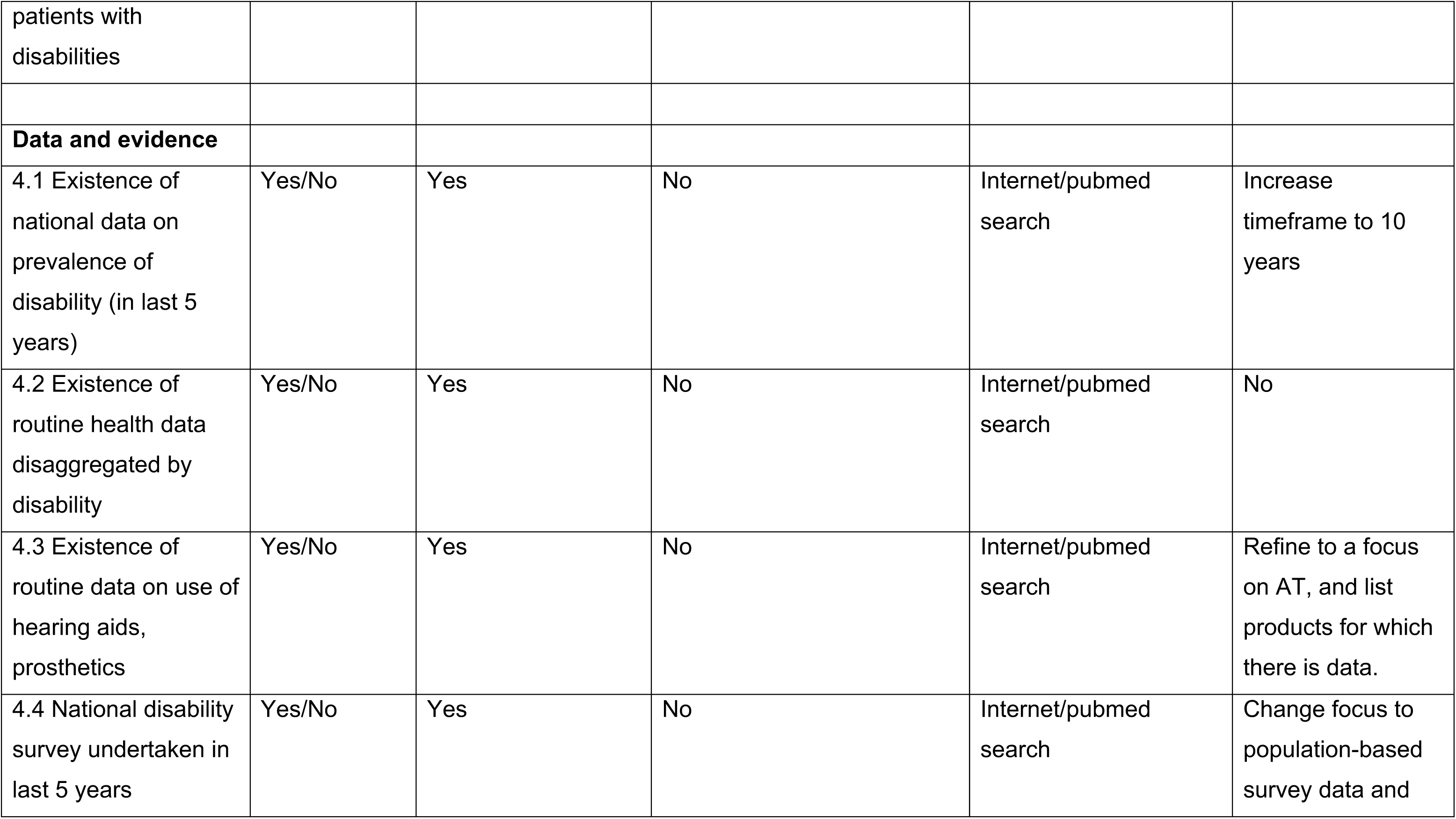
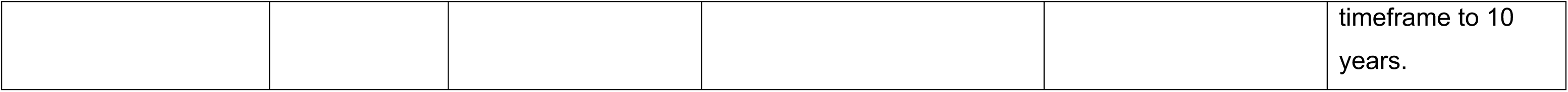
Systems levels indicators, definition and status in Maldives and Zimbabwe.

Status in the Maldives and Zimbabwe: Across both settings, governance appeared broadly supportive of the inclusion of people with disabilities in the health system, including the presence of laws, health sector plans, and ratification of the UNCRPD. However, although, the health sector plans in both Maldives and Zimbabwe mention disability, it is in brief and mostly with respect to preventing disability rather than promoting a disability-inclusive health system. Similarly, the Maldives HIV plan makes brief reference to disability, while the Zimbabwe HIV plan only recognises people with disabilities as a key risk population.

##### 3.2.1.2. Leadership

Four indicators focus on leadership (Table 2). The three indicators related to representation of disability in MoH, national health sector coordination and pandemic preparedness (2.1, 2.2, and 2.4) required clarification on what “responsibility” or “representation” would entail. The indicator on representation in Global Fund Country Coordinating Mechanism was perceived to be clear (2.3), although potentially not relevant in all countries. Most of the data could be gathered through interviews with key stakeholders.

Status in the Maldives and Zimbabwe: There was a general lack of leadership on disability in the health system in the Maldives. In Zimbabwe, there is a MoH department responsible for disability and rehabilitation, and a Disability Board is in place that advises the government, though not specifically on health.

##### 3.2.1.3. Health financing

Three indicators describe health financing, assessing funding for AT/rehabilitation (3.1), budget for disability (3.2), and reimbursement adjustment (3.3)(Table 2).

Broadly, these three indicators were considered adequate, although they would benefit from further refinement (e.g. estimation of funding for AT from other ministries, indication of national or federal budgets, description of reimbursement adjustment mechanisms). The information was obtained through key informant interviews.

Status in the Maldives and Zimbabwe: Funding was allocated for AT/rehabilitation and for disability-related activities in both the Maldives (although not in the MoH budget) and Zimbabwe. The budget amounts were not available to the researchers. The researcher suggested that it may be available through the Ministry of Health, although it may be complex to obtain if the budget is divided across different lines (e.g. sign language interpretation, accessible equipment, transport support).

Reimbursement adjustment occurred in the Maldives through the National Social Protection Agency rather than the MoH. These adjustments were not made in Zimbabwe.

##### 3.2.1.4. Data and Evidence

Four indicators relate to availability of data and evidence, and measure existence of national data or a national survey on disability (4.1 and 4.4) and routine health data on disability (4.2) or AT (4.3) (Table 2). The indicators were perceived to be clear and feasible to collect, with the only refinement being to expand the time frame from 5 to 10 years. The information was obtained through searches of the Internet and scientific literature through Pubmed.

Status in the Maldives and Zimbabwe: There was good availability of data and evidence in the Maldives, including from a national survey of disability conducted in 2017 (Banks et al., 2020). The Maldivian health system operates through a national health insurance programme, which collects electronic data. This electronic data could potentially be linked to disability allowance data to allow disaggregation of health information by disability. It would also allow reporting of use of AT and other disability-relevant care (e.g. physiotherapy). In contrast, there was little data or evidence related to the health needs/coverage of people with disabilities from Zimbabwe. A national survey had been undertaken in 2013, but it was more than five years before the assessment and so was not eligible, although it did provide data on the prevalence of disability and access to health and rehabilitation ( Eide et al., 2013). The national census of 2022 was the first to include disability-related metrics. At the time of the writing of this article, the full report had not been issued.

#### 3.2.2. Service level indicators: Demand

##### 3.2.2.1. Autonomy and awareness

The three indicators related to autonomy and awareness focussed on whether Organizations of Persons with Disabilities (OPDs) were active in advocating about health (5.1), evidence of autonomy and awareness for people with disabilities (5.2), and availability of health information in accessible formats (5.3) (Table 3). The group concluded that some refinement of the indicators was needed. For example, there are often too many OPDs in most countries to allow assessment of all of their activities, and the indicator does not specify the type or intensity of advocacy.

**Table 3:** Service levels indicators, definition and status in Maldives and Zimbabwe.

| Indicator | Measure | Maldives status | Zimbabwe status | Source of information | Revision needed |
| --- | --- | --- | --- | --- | --- |
| **Demand: Autonomy and awareness** |  |  |  |  |  |
| 5.1 Organizations of Persons with Disabilities (OPDs) advocate for healthcare access for people with disabilities | Yes/No | Yes | Yes | Key informant interview(s) and internet search | Focus on lead national OPDs and clarification on level of activity |
| 5.2 Persons with disabilities report autonomy and awareness about health access | Yes/no and descriptor | Yes, national survey | Yes, national survey | PubMed and internet search | Limit to surveys comparing people with and without disabilities in terms of autonomy and awareness |
| 5.3 Key health information is available in accessible formats | % of clinics that offer alternative communication methods | Not available | Not available | Key informant interview(s), PubMed and internet search | Adapt measure as it is too specific and unlikely to be available. |
| **Demand:**<br>**Affordability** |  |  |  |  |  |
| 6.1 Disability allowance available in country | Yes/No | Yes | No | Key informant interview(s), and policy review | Include descriptor on entitlements and coverage |
| 6.2 Transport subsidies are available for people with disabilities | Yes/No | Yes | No | Key informant interview(s), and policy review | No change needed |
| 6.3 Health insurance coverage for people with disabilities | Yes/No | Yes | No | Key informant interview(s), and policy review | Clarification of health coverage not only through health insurance, but also other schemes (e.g. disability allowance) |
| 6.4 Availability of out-of-pocket healthcare spending, disaggregated by disability | \$ per year, by disability status | \$7.8 for people with disabilities and \$4.1 for people without disabilities (but national health insurance in place for all) | Data not available. | Internet and PubMed search | Exclude, as data not available disaggregated by disability in most settings. |
| 6.5 Household spending on health, disaggregated by disability | % of household expenditure on health as share of total household expenditure/income, by disability status | 0.4% for people with disabilities and 0.2% for people without disabilities (but health insurance in place for all) | Data not available | Internet and PubMed search | Exclude, as data not available disaggregated by disability in most settings. |
| **Supply: Human resources** |  |  |  |  |  |
| 7.1 Curricula for medical doctors include mandatory disability element | Number of hours training | Data not available | No | Key informant interview(s) | Rephrase as yes/no and number of hours, and focus on national curriculum |
| 7.2 Curricula for nurses include mandatory disability element | Number of hours training | Data not available | Yes; amount varies by nursing school | Key informant interview(s) | Rephrase as yes/no and number of hours, and focus on national curriculum |
| 7.3 Curricula for community health workers (CHW) include mandatory disability element | Number of hours training | Data not available | Yes; CHW curriculum includes some training on disability | Key informant interview(s) | Rephrase as yes/no and number of hours, and focus on national curriculum |
| 7.4 People with disabilities are represented in the health workforce | % of health workforce with disability | Data not available | Data not available | Internet search | Focus on % of medical doctors that have a disability |
| 7.5 People with disabilities report that they feel well treated by health workers | % respondents | People with disabilities reported more negative attitudes from staff when accessing health services than people without disabilities (15% versus 8%). | Data on lack of satisfaction from national survey. | PubMed and internet search | Limit to quantitative survey comparing people with and without disabilities |
| **Supply: Health facilities** |  |  |  |  |  |
| 8.1 Existence of national accessibility standards established | Yes/no | No | No | Internet search, key informant interview(s) | No change needed |
| 8.2 Accessibility of facilities | % of facilities that meet national standard | No national standard or data on accessibility | No national standard or data on accessibility | PubMed search | Adjust to whether an accessibility audit had been undertaken in last 10 years. |
| **Supply: Specialized services and AT** |  |  |  |  |  |
| 9.1 National law and/or policy on specialist services and AT | Yes/no | Yes | No | Internet search | Overlaps with data collected under governance. |
| 9.2 National assessment on AT and/or rehab done in last 5 years | Yes/No | No | No | Internet search | Extend to within last 10 years |
| 9.3 Coordination mechanism for specialist services and AT | Yes/no | No | Yes; Rehabilitation department in MoH | Key informant interview(s) | Clarify that this is a coordination mechanism across<br>ministries |
| 9.4 Trained workforce<br>available to provide<br>specialist services<br>and/or AT | Number of<br>physiotherapists per<br>10,000 population | Data lacking | Data lacking | Internet and PubMed<br>search | No change<br>needed |

Quantitative data was available in the Maldives and Zimbabwe from the national surveys on autonomy and awareness comparing people with and without disabilities (Banks et al., 2020; Eide et al., 2013). However, in other settings it is likely that only qualitative evidence will be available with a lack of comparable data for people without disabilities, making it difficult to make inferences about inequities. The measure for the indicator “key health information is available in accessible formats” related to the proportion of clinics that offer alternative communication methods, and this was deemed too specific and unlikely to be available in most settings. The information for this indicator set was obtained through a combination of key informant interviews, internet searches and searches of the scientific literature.

Status in the Maldives and Zimbabwe: There was evidence of OPDs advocating for healthcare access for people with disabilities in both countries, although it was believed that it was unlikely to be a priority for many OPDs. The Maldives national survey (2017) documented widespread barriers that people with disabilities faced in accessing healthcare services compared to people without disabilities (Banks et al., 2020). The Zimbabwe survey (2013) also showed issues related to autonomy facing people with disabilities in terms of unmet healthcare needs, problems understanding health information and barriers to care seeking (Eide et al., 2013).

##### 3.2.2.2. Affordability

Affordability of healthcare by people with disabilities, was assessed in terms of availability of disability allowance (6.1), transport subsidies (6.2) and health insurance (6.3) and evidence on health expenditure by disability status (out of pocket: 6.4, household spending: 6.5) (Table 3). These indicators were reported to be clear and easy to collect, although further clarifications may be helpful (e.g. reporting coverage of the disability allowance among people with disabilities). Data on out-of-pocket expenditure or household expenditure on health disaggregated by disability was available for the Maldives, but not Zimbabwe (Banks et al., 2020). We considered it unusual for this data to be available, and the recommendation was to drop this indicator. An identified gap where a new indicator would be helpful was the presence of “co-payments” or any additional charges not covered by the insurance or the government for services being waived for people with disabilities. This information was largely obtained through key informant interviews, although there were difficulties gaining access to relevant stakeholders in Zimbabwe.

Status in the Maldives and Zimbabwe: Efforts to improve affordability of healthcare for people with disabilities were in place in the Maldives, including provision of a disability allowance, transport subsidies and health insurance, while these were lacking in Zimbabwe (Hameed et al., 2022a). In the Maldives, health expenditure (both absolute or as a proportion of household income) was twice as high for people with disabilities as compared to those without, although the absolute levels were very low as there is a national health insurance programme in place (unpublished data).

#### 3.2.3. Service level indicators: Supply

##### 3.2.3.1. Human Resources

Three indicators assessed inclusion of disability in medical curriculum for different cadres (7.1 – 7.3), and the remainder assessing representation of disability in the health workforce (7.4) and reported discrimination by people with disability (7.5) (Table 3). Data was lacking in the Maldives as to whether disability was included in healthcare worker curricula, and in Zimbabwe information on the number of hours trained was not available. Data was lacking on the proportion of the health workforce that have a disability in both countries. The suggestion is to focus this latter indicator on doctors and nurses specifically, and separately, as these are relatively internationally consistent cadres that may have available recorded data {Association, 2020 #1451}. The indicator on whether people with disabilities report being well-treated by health workers should be restricted to quantitative data where there is a comparison of people with and without disabilities. This information was largely obtained through key informant interviews, review of national reports and a search of the scientific literature through Pubmed.

Status in Maldives and Zimbabwe: For the Maldives, data was only available for the healthcare worker attitude indicator, which showed that people with disabilities were more likely to report negative attitudes from staff when accessing healthcare services (15%) in comparison to people without disabilities (8%) (unpublished data from national survey) (Banks et al., 2020). In Zimbabwe, nurses received training on disability, although doctors did not. Community health workers received training on disability in specific topics of the curriculum (e.g. home-based care, pregnancy and patient management). The national survey in Zimbabwe showed that negative attitudes and prejudice were an issue facing people with disabilities, but this was not measured specifically with respect to healthcare (Eide et al., 2013). Qualitative research demonstrated that people with disabilities experienced negative attitudes from healthcare workers (Smythe et al., 2022a, b).

##### 3.2.3.2. Health facilities

The indicator on the existence of national accessibility standards (8.1) was perceived to be clear and relevant (Table 3). The second indicator considered the proportion of facilities that met national standards and was not relevant given the lack of these standards (8.2). Consequently, it was recommended to change this indicator to focus on whether an accessibility audit had been undertaken in the last decade. We sought this evidence from key informant interviews, a review of national reports and a search of the scientific literature through Pubmed.

Status in the Maldives and Zimbabwe: No data was available in either county for these indicators.

##### 3.2.3.3. Specialized services and AT

The first indicator related to whether a national law or policy existed on these services (9.1), and the decision was made to move this indicator into the governance section (Table 3). The remaining three indicators focussed on national assessment of AT and/or rehabilitation (9.2), existence of a coordination mechanism (9.3) and the scale of the relevant workforce (9.4). These indicators were perceived to be clear and required only minor clarifications. The reviewers suggested to rephrase this component as “Rehabilitation services and AT”, to emphasise that rehabilitation should be part of routine care and not something “special”. This information was largely obtained through key informant interviews and a review of national reports.

Status in the Maldives and Zimbabwe: In the Maldives, a goal existed for specialist services/AT in the national health policy and in Zimbabwe there was a coordination mechanism for these services. Otherwise, the status was poor in both countries for these indicators. Data were lacking on the number of physiotherapists per 10,000 population, although this information could potentially be obtained through the MoH.

#### 3.2.4. Outputs and Outcomes

The seven output indicators and seven outcome indicators are consistent with the priorities of UHC monitoring. For outputs, these are comparisons of people with and without disabilities in terms of: Women whose demand is satisfied by modern contraception (10.1); People with HIV receiving ART (10.2); Children aged 12-23 months who have received DTP3 (10.3); People with refractive error who have glasses (10.4); People with diabetes on treatment (10.5); People with hypertension on treatment (10.6); Women receiving breast cancer screening (10.7). Outcome measures were the following, disaggregated by disability: Under 5 mortality rate (11.1); Overall mortality rate (11.2); Prevalence of diabetes among people aged 18+ (11.3); Prevalence of HIV (11.4); Prevalence of overweight and obesity in people aged 18+ (11.5); Prevalence of children wasted aged 0-59 months (11.6); Prevalence of hypertension in people aged 18+ (11.7). These indicators were perceived to be clear. However, the data was largely not available disaggregated by disability in either the Maldives or Zimbabwe. A recommendation was made to replace these indicators with those derived from widely collected data, namely from the UNICEF Multiple Indicator Cluster Surveys (MICS) and Demographic and Health Surveys (DHS) (i.e. modern contraceptive coverage, ART coverage, DTP3 coverage, refractive error coverage, NCD coverage, mortality, and prevalence of diabetes, HIV, overweight/obesity and wasting – detailed description in Appendix). This information was largely sought through a search of the scientific literature using Pubmed.

Status in the Maldives and Zimbabwe: Evidence from the Maldives showed that people with disabilities were more likely to have a diagnosis of diabetes or hypertension than people without disabilities (17% versus 12%; 34% versus 21%) (unpublished data from national survey) (Banks et al., 2020), but that there was no difference in treatment levels for these conditions. The data exists for the remaining indicators in the Maldives, but had not been analysed or reported disaggregated by disability. In Zimbabwe, data were not available for the majority of indicators, excepting prevalence of diabetes and hypertension by disability status, both of which conditions were more prevalent in people with disabilities compared to those without (hypertension: 11.1% versus 2.4%; diabetes: 2.3% versus 0.3%) (Eide et al., 2013).

Revised indicators and definitions are presented in Web Appendix Table.

### 3.3. Feedback on indicator set application

The primary objectives of the indicator set were determined by consensus and include to: 1) collate data on the level of disability inclusion in the health system to set a benchmark, 2) identify ways in which the health system could be improved to become more disability-inclusive, and 3) generate monitoring data on change in disability inclusion through repeated use. It was agreed that the application of the indicator set should be led by the MoH, or by a consultant (e.g. NGO) working in collaboration with the MoH. People with disabilities should be active partners in this process. Additional users of the indicator set could include researchers to undertake situational analyses or to identify indicators to use in studies, disability programs to monitor inclusion and disability rights groups to identify gaps and advocate for action.

Key identified challenges were that a disability focal person may be lacking at the MoH, resulting in limited drive of the process. Furthermore, MoH partners may consider that disability is the responsibility of other departments (e.g. Ministry of Social Welfare), and indeed a plan for disability-inclusive health may need the involvement of a range of Ministries. The assessment may need to include other key partners, such as the Ministry of Finance or Social Welfare. Another concern was that the MoH may be reluctant to reveal poor levels of disability inclusion and thus be reticent to undertake the assessment using the indicator set. Education and advocacy are therefore important to demonstrate the importance of disability-inclusive health, including the financial benefits.

In terms of logistics, it was anticipated that the timeline for the assessment would be 2-3 months. Provision of tools (e.g. formatted excel worksheets) and good practice examples of use would assist with scale up of the use of the indicator set. It was advised that the assessment should commence with an inception workshop attended by key partners (e.g. MoH, OPDs, relevant NGOs, healthcare workers, representatives from other ministries). After the assessment, there should be a sharing and strategic planning workshop to agree key recommendations and actions. This workshop should be attended by MoH, people with disabilities and other key audiences (e.g. relevant NGOs and OPDs who can support action and hold the MoH to account). Developing and sharing examples of disability strategies and good practice could help in plan development to implement the agreed recommendations and actions. Furthermore, there should be clear plans for costing and accountability (e.g. monitoring of implementation).

A website was created describing the assessment purpose, process and indicators: https://www.themissingbillion.org/assessment-toolkit.

## 4. Discussion

### 4.1 Key findings

There is growing awareness of the importance of promoting disability-inclusive health systems, as they will help to achieve the right to healthcare for people with disabilities (UN, 2006), and improve their health outcomes (Kuper & Heydt, 2019; WHO, 2011, 2022). Disability-inclusive health systems are also likely to be more resilient and work better for all (e.g. minority language speakers, people with temporary impairments), and are expected be cost-saving (WHO, 2022). Currently, few global actors focus on disability-inclusion in their programmes. Key reasons are a lack of awareness of the issue, little knowledge of where the gaps are in their activities, and a lack of clarity on the priority areas for intervention. The Missing Billion Disability-Inclusive Health System framework supports a structured approach to assessing the inclusion of people with disabilities holistically by health systems, with specific indicators related to different components. This assessment approach was considered by international key informants to be logical and comprehensive.

The pilot data from the Maldives and Zimbabwe demonstrated that it was feasible to collect indicators related to the framework components to indicate the level of disability-inclusion in the health system. The indicators were viewed as mostly useful and relevant, with some requiring refinement, usually to improve clarity. Moreover, the assessment revealed areas where the health system in the Maldives and Zimbabwe was making progress in terms of disability inclusion (e.g. in terms of legal framework) and other areas where there were large gaps (e.g. leadership on disability) or lack of data (e.g. accessibility, outputs and outcomes).

### 4.2. Implications for policy and practice

The key potential value of the indicator set is to collate data on the level of disability inclusion in the health system to set a benchmark and help to identify ways in which the health system could be improved to be more disability-inclusive. It will also allow international comparison to reveal good practice (Bitton et al., 2017; Fekri et al., 2018), and produce data on change in disability inclusion through repeated use.

Existing tools (e.g. PHCPI) may show overall health system performance, but not whether achievements are equitable and reach all segments of the population unless special attention is provided. As indicated already, people with disabilities will need additional focus as they face additional barriers to accessing services, which is in violation of their rights as set out in UNCRPD. The strength of the indicator set is therefore that it fills an important gap, as there are no other comprehensive health system performance assessment tools that have a specific focus on disability-inclusion. Indeed, it has already been used to inform guidance on World Bank health care investments to promote disability-inclusion (World Bank, 2022), as well as a WHO Euro policy brief on disability-inclusive health systems (WHO, 2021b). Furthermore, the indicator set fulfils many of the criteria of a ‘good’ health system performance assessment tool; it was developed in a structured and inclusive way, based upon a conceptual framework, addresses an important policy issue, and presents clear constructs and indicators (Tashobya et al., 2014).

A key concern is that there is not yet a structure in place to promote scale-up of the assessment using the indicator set (e.g. incorporation in World Bank or WHO strategy) (Tashobya et al., 2014). The success of the indicator set is therefore reliant on buy-in and implementation by individual MoHs, working in collaboration with people with disabilities (e.g. OPDs) (Tashobya et al., 2014). However, currently many MoHs lack capacity, are under-funded and perceives disability as a low priority. Advocacy is therefore needed to raise the priority of this issue, or the availability of seed funding or technical assistance. Incorporating the indicator set with existing tools may also encourage uptake, whether general health systems assessments (e.g. PHCPI, UHC monitoring initiatives), or disability-relevant assessments (e.g. WHO Systematic Assessment of Rehabilitation Situation) (Kleinitz et al., 2022), and we attempted to produce an indicator set which was complementary to these approaches. Furthermore, countries may be cautious about undertaking such an assessment if they perceive their country to be under-performing or if the resources and political will are not available to be able to respond to identified gaps. Care will therefore need to be taken on how to describe the purpose of the assessment, and ensure that this is viewed as an opportunity for measurement and improvement, rather than judgment. The indicator set may need to be tailored to different settings, including additional indicators viewed to be relevant or important for that setting (Ratcliffe et al., 2019). Another issue is that donors are generally not yet sufficiently committed to disability inclusion to encourage programmes or governments to make health systems disability-inclusive. This situation may be changing; The FCDO disability inclusion and rights strategy 2022 to 2030 makes commitments to “Achieving inclusive health for all” (FCDO, 2022), and the World Bank has recently published guidance on creating disability-inclusive health care systems (World Bank, 2022).

There are several limitations to the indicator set which need consideration. The pilot study in the Maldives and Zimbabwe showed that there was a lack of available data for several of the indicators, although this gap may be filled if the assessment is led by the MoH as they will have access to unpublished data. Indeed, the lack of involvement of the MoH in the pilot study is an important limitation. The indicator set focuses on measures within the health system, to drive action by health actors. Yet, several important actors for disability-inclusive health are outside of the health system, such as disability allowance providers and programmes to provide AT (Hameed et al., 2022b). Social determinants of health are important, perhaps particularly for people with disabilities, yet lie outside the boundary of the health system. The indicator set also does not focus on broader societal goals that are important for people with disabilities, such as increasing employment. We also did not undertake psychometric assessment of the indicator set. The assessment process was undertaken within the context of an ongoing research study in both settings. Consequently, we could not measure the time taken for the assessment alone. Moreover, this set-up may mean that the assessment appeared more feasible than it would in other settings, potentially limiting generalizability. Feasibility of assessment implementation was considered qualitatively, rather than through the use of objective measures. Finally, the Missing Billion Disability-Inclusive Framework was produced through adaptation from two existing frameworks, and not developed through new conceptual or theoretical work.

### 4.3. Future work

The indicator set needs to be implemented in more countries to allow a fuller assessment of feasibility (Bowen et al., 2009), including with objective measures of feasibility. Wider use will also allow finalisation of the indicator set, for instance, determining whether additional indices are needed, more response levels required, and composite indices should be created (e.g. for the outcomes and outputs).

Further consideration is also required of what is needed for the assessment to guide policy and programmatic improvement. A key priority is to encourage ownership and involvement by the MoH, and to generate and share good practice in the application of the indicator set. Consideration of how the international data is collated and displayed is also important, potentially through data visualization tools such as used by PHCPI (PHCPI, 2018). International comparison can generate sensitivity, as it implies judgement, and may also require a mechanism to review and validate results before they are displayed. The indicator set can also be adapted to assess disability-inclusion in specific aspects of care, such as in HIV programmes.

### 4.4. Conclusions

Health systems have neglected the inclusion of people with disabilities, leading to a violation of their rights and worse health access and outcomes. This new assessment approach can help to identify key issues and guide action, and thereby may ultimately improve health systems for all.

## Data Availability

All data produced in the present study are available upon reasonable request to the authors

## Acknowledgements

We are grateful to Scope International in their support in the refinement of the indicator set.

