## Supplement table for "The process of developing and piloting a tool in the Maldives and Zimbabwe for assessing disability inclusion in health systems performance"

Web Appendix Table: Revised indicator list and definition

|  | **Indicator** | **Definition** | **Metric** |
| --- | --- | --- | --- |
|  | **System: Governance** |  |  |
| 1.1 | UNCRPD | Ratification of UNCRPD by country | Yes/No |
| 1.2 | National law | Existence of a national law protecting the rights of persons with disabilities to health | Yes/No  Description of whether:   - law prohibits discrimination - law requires reasonable accommodation - law is disability-focused or health focused |
| 1.3 | National health policy | Existence of a national policy or decree on health for persons with disabilities | Yes/No  Description of whether the following are specified:   - how policy ensures access to specialist (rehabilitation services, AT) and general healthcare services for persons with disabilities |
| 1.4 | National Health Sector Plan(s) | Inclusion of people with disabilities in National Health Sector Plan(s) | Yes/No and description  Description of whether the following are specified:   - actions and targets for specialist health services for persons with disabilities - actions and targets for general health care for persons with disabilities (not prevention of disability) - basic statistics provided about persons with disabilities and health - monitoring and evaluation indicators on disability as part of overall framework for the health sector |
| 1.5 | National HIV plan | Inclusion of people with disabilities in National HIV plan | Yes/No and description  Description to include:  1) Includes actions and targets for specialist HIV health services for persons with disabilities 2) Includes actions and targets for general HIV health care for persons with disabilities (not only prevention of disability) 3) Includes basic statistics about persons with disabilities and HIV 4) Includes monitoring and evaluation indicators on HIV amongst persons with disabilities as part of overall framework for the health sector |
| 2.1 | MoH leadership | Existence of a focal point/team/directorate in MoH that’s responsible for ensuring health access for people with disabilities | Yes, responsibility for disability inclusion, and title of role/team  Yes, responsibility for rehabilitation, and title of role/team No, neither |
| 2.2 | National health sector coordination | Formal representation of persons with disabilities (individual, or OPDs) in highest-level health sector coordination structure | Yes, and title of structure/group No |
| 2.3 | Global Fund CCM | Representation of person with disabilities in Global Fund CCM | Yes/No |
| 2.4 | Pandemic preparedness structures | Formal representation of people with disabilities (individuals are representing OPD) in national COVID-taskforce | Yes/No |
| 3.1 | AT/rehabilitation budget | Funding for AT/rehabilitation in MoH (or devolved levels) budget | % of annual MoH budget, or absolute amount contributed from other Ministries as % MoH budget |
| 3.2 | Disability inclusion budget | Budget (MoH or devolved levels) for role/department in MoH working on disability inclusion | Yes, at the federal and indication of $ amount Yes, at the decentralized level and indication of $ amount for one example location No, neither |
| 3.3 | Reimbursements | Reimbursement adjustment for services provided to patients with disabilities | Yes, there is a national health insurance and reimbursement for at least certain conditions/treatments to health providers is adjusted for disabilities  Yes, there are private insurances and reimbursement for at least certain conditions/treatments to health providers is adjusted for disabilities  Yes, there is a national taxation-based budget and any capitation rates for devolved levels is adjusted for disabilities  No adjustments (with whatever financing mechanism) |
| 4.1 | Prevalence of disability | Existence of national data on prevalence of disability (from within last 10 years) | Yes/No |
| 4.2 | Routine health data | Existence of routine health data disaggregated by disability | Yes/No |
| 4.3 | AT coverage | Existence of data for coverage (% of need covered) of AT | Yes and list of products for which there is data No |
| 4.4 | Population-based data on disability and health | National disability survey done in last 10 years (including health data) | Yes/No |
| 5.1 | OPD advocacy | OPDs advocate on the right to health for persons with disabilities with government and NGO delivery partners | Yes - website of 1-3 national leading OPDs describe priorities/programs/work on advocacy on health on website |
| 5.2 | Autonomy and awareness | People with disabilities report autonomy and awareness about health access | Yes - in a quantitative survey from within the last 10 years persons with disabilities were asked about autonomy and awareness about health (in comparison to people without disabilities) OR Yes - qualitative data published in the last 10 years in a peer-reviewed journal on reported autonomy and awareness about health |
| 5.3 | Accessibility of health information | Health information is available in accessible formats | Yes - national COVID information and announcement made available at least in the 2 out of the 4 following alternative formats: simple language, sign interpretation of video/tv messages, braille, information for care givers |
| 6.1 | Disability allowance | There is a disability allowance that is available to cover healthcare fees not covered by existing insurance or tax-based systems, e.g. travel to clinics, AT | Yes - and % of population covered, amount per person per time unit in $ No |
| 6.2 | Transport subsidy available for disabled people | Transport subsidy is available | Yes/No |
| 6.3 | Health coverage | People with disabilities are fully covered for free healthcare through social health insurance, tax-based system, provision as part of disability allowance or any other stipulations | Yes/No |
| 6.4 | Co-pays | Any co-pays for services in either health insurance or taxation based systems are waved for persons with disabilities | Yes/No |
| 7.1 | Training of medical doctors | Information about disability delivered as part of the national curricula for medical schools/colleges | Yes - number of hours training  No |
| 7.2 | Training of nurses | Information about disability delivered as part of the national curricula for nurses/nursing colleges | Yes - Number of hours training  No |
| 7.3 | Training of CHWs | Information about disability delivered as part of the national CHW training curricula | Yes - Number of hours training  No |
| 7.4 | Representation in health workforce | People with disabilities are represented in the health workforce | % of medical doctors that have disability |
| 7.5 | Satisfaction | People with disabilities report that they feel well treated by health workers | Yes - in a quantitative survey from within the last 10 years persons with disabilities were asked about satisfaction with health worker services (in comparison to people without disabilities) OR Yes - qualitative data published in the last 10 years in a peer-reviewed journal on reported satisfaction |
| 8.1 | National accessibility standards | Existence of national accessibility standards | Yes/No |
| 8.2 | Accessibility of facilities | Accessibility audit of health facilities has been undertaken in the last 10 years | Yes - results of audit report in published government report/documents or peer-reviewed journal No |
| 9.1 | National assessments | National assessment on AT or rehabilitation (e.g. STAR or RATA) done in the last 10 years | Yes/No |
| 9.2 | Coordination | Coordination mechanism cross-Ministry for rehabilitation services and AT where more than 1 ministries involved | Yes No  No - because only 1 ministry responsible for AT/rehabilitation |
| 9.3 | Trained workforce available to provide rehabilitation services and AT |  | # of Physiotherapists/10,000 population |
| 10.1 | Modern contraception coverage | Women whose demand is satisfied with a modern method of contraception, disaggregated by disability | % of women with disabilities, compared to % of overall women |
| 10.2 | ARTs coverage | People with HIV receiving ART, disaggregated by disability | % of people with disabilities that have coverage, compared to coverage of people without disabilities |
| 10.3 | DTP 3 coverage | Children aged 12-23 months who have received diphtheria-tetanus-pertussis vaccine (DTP3), disaggregated by disability | % of children with disabilities, compared to % of overall children |
| 10.4 | Refractive error coverage | People with refractive error have coverage of glasses | % of those with need who have glasses (e.g. from RAAB survey) |
| 10.5 | NCD coverage | People with diabetes on treatment OR people with hypertension on treatment, disaggregated by disability | % of people with disabilities, compared to people without disabilities |
| 11.1 | Mortality | Overall mortality rate, disaggregated by disability | Deaths per 100 000 population; people with disabilities compared to people without disabilities |
| 11.3 | Diabetes | Prevalence of diabetes OR hypertension among persons aged 18+ years, disaggregated by disability (Global Monitoring Framework NCDs; indicator #12, indicator #11, WHO) | People with disabilities, compared to people without disabilities |
| 11.4 | HIV | Prevalence of HIV, disaggregated by disability | % of people living with HIV among adults aged 15–49; people with disabilities compared to people without disabilities |
| 11.5 | Overweight and obesity | Prevalence of overweight and obesity among persons aged 18+ years, disaggregated by disability (Global Monitoring Framework NCDs; indicator #13, WHO) | % of all population with disabilities, compared to population without |
| 11.6 | Wasting | Prevalence of children wasted (moderate and severe), 0-59 months of age, disaggregated by disability; WHO Child Growth Standards median | % of children with disabilities, compared to % of overall children |
